# Multi-trait Polygenic Profiling and Survival Free of Dementia and Disability: Results from the Health and Retirement Study

**DOI:** 10.64898/2026.09.09.26362686

**Authors:** Yome Tawaldemedhen, Santiago Clocchiatti-Tuozzo, Cyprien Rivier, Shufan Huo, Andrew Silberfeld, Tim D’Aoust, Stephanie Debette, Adam de Havenon, Thomas M. Gill, Guido J. Falcone

## Abstract

**Objective:** To determine whether a multi-trait polygenic profile for dementia is associated with dementia- and disability-free survival in middle-aged and older adults.

**Background:** Clinical trials of brain health and aging increasingly use holistic, patient-centered outcomes that capture overall health status. Survival free of dementia and disability is one such outcome. We hypothesized that adverse polygenic profiles for dementia would be associated with higher risk of dementia, disability, or death.

**Design/Methods:** We conducted a genetic association study within the Health and Retirement Study, a prospective, nationally representative longitudinal cohort of U.S. adults. Polygenic profiling used the previously validated integrated polygenic risk score for dementia (iPRS-DEM), which incorporates neurodegenerative and vascular genetic components. Participants were categorized as having favorable (≤20th percentile), intermediate (20th-80th percentile), or poor (>80th percentile) polygenic profiles. The primary outcome was a composite of dementia, disability, or death. Cox proportional hazards models were adjusted for age, sex, genetic ancestry, and genetic principal components. Interaction between continuous iPRS-DEM and APOE ε4 status was assessed for 31-year outcome risk.

**Results:** Among 45,234 HRS participants, 15,620 provided DNA samples, 15,565 had genotype data after quality control and imputation, and 14,333 were free of dementia and disability at baseline (median age 55 years; 58% female). Compared with a favorable polygenic profile, intermediate and poor profiles were associated with higher risk of the composite outcome (HR 1.14, 95% CI 1.07-1.21 and HR 1.43, 95% CI 1.26-1.63, respectively). The relative association was strongest for dementia; for poor versus favorable profiles, HRs were 1.58 (95% CI 1.21-2.05) for dementia, 1.43 (95% CI 1.14-1.79) for disability, and 1.33 (95% CI 1.15-1.54) for death. An interaction between continuous iPRS-DEM and APOE ε4 status was observed for 31-year dementia risk (P=0.016). Relative to favorable-profile APOE ε4 non-carriers, dementia risk was highest among participants with a poor profile who were APOE ε4 carriers (HR 2.32, 95% CI 1.73-3.12).

**Conclusions:** Adverse polygenic profiles for dementia were associated with higher composite risk of dementia, disability, or death in a large population-based cohort, with the strongest association observed for dementia. Joint consideration of iPRS-DEM and APOE ε4 further identified individuals at elevated dementia risk. These findings support the potential value of polygenic profiling for risk stratification while highlighting limitations related to ancestry, generalizability, and clinical translation.

## INTRODUCTION

As life expectancy continues to rise, preserving cognitive and physical independence in older adults has become a central goal of modern medicine. This shift from extending lifespan to promoting health span has motivated the use of composite outcomes that capture multiple domains of well-being in aging. Among these, survival free of dementia and disability has emerged as a particularly meaningful endpoint, integrating cognitive and functional dimensions that reflect overall health status rather than the absence of disease alone.^1^ Large pragmatic trials, such as the Pragmatic Evaluation of Events and Benefits of Lipid-Lowering in Older Adults (PREVENTABLE) and the Statins in Reducing Events in the Elderly (STAREE), have adopted this outcome to evaluate interventions that may extend healthy aging, underscoring a growing emphasis on patient-centered measures of longevity.^2,3^

Genetic variation contributes substantially to inter-individual differences in late-life trajectories, influencing susceptibility to both neurodegenerative and vascular conditions. APOE remains the most established genetic determinant of aging-related outcomes. The ε4 allele increases the risk of Alzheimer disease, vascular disease, and mortality, whereas the ε2 allele confers relative protection across several phenotypes.^4–7^ Recent analyses from the Health and Retirement Study (HRS) demonstrated that APOE ε4 variants are also associated with a composite outcome of dementia, disability, and death.^8^ These findings highlight the potential of integrating genomic information into clinical trials of older adults, where genetic heterogeneity may modify both absolute and relative treatment effects.

While APOE captures a portion of genetic susceptibility, dementia and disability arise from complex interactions between neurodegenerative and vascular pathways. Building on this concept, the integrated polygenic risk score for dementia (iPRS-DEM) was developed to aggregate genetic signals from Alzheimer disease and 23 additional vascular or neurodegenerative traits, providing a multidimensional representation of dementia-related biology.^9^ The iPRS-DEM predicts dementia independently of APOE and demonstrates strong performance in European and East Asian ancestry groups; however, validation in African or Hispanic ancestry participants was not statistically significant despite directionally consistent estimates.^9^ These findings emphasize both the translational potential and the ancestry-related limitations of current polygenic models.

To address these knowledge gaps, we examined whether a polygenic profile encompassing neurodegenerative and vascular risk was associated with the risk of dementia, disability, or death in the HRS. We further assessed whether combining iPRS-DEM categories with APOE ε4 status enhances genetic risk stratification and whether these associations can be reproduced in both European and African ancestry groups, recognizing that iPRS-DEM was originally derived in European ancestry cohorts and that polygenic score distributions often differ across ancestry groups.^9–12^

We hypothesized that individuals with unfavorable polygenic profiles would experience a higher risk of dementia, disability, or death, and that joint classification of iPRS-DEM and APOE ε4 status would identify subgroups at particularly elevated risk. By integrating comprehensive polygenic modeling with a patient-centered measure of aging, this study seeks to extend genomic risk prediction to outcomes that reflect functional health and to evaluate its applicability across ancestries in the context of precision prevention and healthy longevity.

## METHODS

### Study design

We conducted a genetic association study nested within the Health and Retirement Study (HRS), an ongoing, nationally representative longitudinal cohort study of adults in the United States designed to investigate health and aging.^13^ The HRS enrolls community-dwelling adults aged 50 years and older and conducts biennial follow-up interviews collecting information on health status, cognitive function, sociodemographic characteristics, and mortality.^13^ For the present study, we included HRS participants with available genetic data and excluded individuals with prevalent dementia or disability at baseline.^13,14^ Baseline was defined as each participant’s entry wave into the HRS, even when genetic sample collection occurred at a later study wave. Participants were followed from baseline until the occurrence of the outcome of interest, loss to follow-up, or the end of the study period, whichever occurred first.

### Genotyping

Genetic data were obtained from participants in the HRS who consented to DNA collection during study waves conducted between 2006 and 2010.^14^ Of the 45,234 individuals enrolled in the HRS cohort, 15,620 provided DNA samples, which were collected using saliva-based methods during enhanced face-to-face interviews.^14^ Genotyping was performed at the Center for Inherited Disease Research using Illumina HumanOmni2.5 arrays, capturing approximately 2.5 million single nucleotide polymorphisms (SNPs).^14^ Genotype calling was conducted using GenomeStudio software, and additional variant calling refinement was performed using zCall to improve detection of heterozygous genotypes.^14,15^ Standard quality control procedures were applied at both the sample and variant levels. Samples with high missing call rates (>2%), chromosomal anomalies, or unexpected relatedness were excluded, and duplicate samples were identified and resolved.^14^ SNPs were filtered based on call rate, Hardy-Weinberg equilibrium, and minor allele frequency thresholds.^14^ Genotype data were pre-phased using SHAPEIT^16^ and imputed to the 1000 Genomes Project reference panel using IMPUTE2.^14,17^ Principal component analysis was performed on genome-wide genotype data to characterize population structure and identify ancestry outliers.^14^ Additional details regarding genotyping and quality control procedures have been described previously.^14^

### Polygenic profile

Polygenic risk was modeled using the integrated polygenic risk score for dementia (iPRS-DEM), previously developed to capture genetic contributions from both neurodegenerative and vascular pathways.^9^ The score integrates genetic variants associated with Alzheimer disease and 23 additional vascular and neurodegenerative traits derived from genome-wide association study summary statistics, with the APOE region excluded from score construction. The iPRS-DEM is publicly available through the Polygenic Score Catalog (PGS ID: PGS005170) and includes approximately 1.32 million genetic variants.^9^

For each participant, the polygenic score was calculated as the weighted sum of risk alleles across included variants and standardized to have a mean of 0 and a standard deviation of 1 within the analytic sample. To facilitate interpretation and evaluate risk stratification, participants were categorized into polygenic profile groups based on the distribution of the score: favorable (≤20th percentile), intermediate (>20th to 80th percentile), and poor (>80th percentile).

### APOE genotype

APOE genotype was determined using the single nucleotide polymorphisms rs429358 and rs7412, which jointly define the three common APOE alleles (ε2, ε3, and ε4).^14^ Participants carrying at least one ε4 allele (ε2/ε4, ε3/ε4, or ε4/ε4) were classified as APOE ε4 carriers, whereas those without an ε4 allele (ε2/ε2, ε2/ε3, or ε3/ε3) were classified as non-carriers.

### Outcome definitions

The primary outcome was a composite measure of dementia, disability, or death. Incident dementia was identified using the validated HRS cognitive classification algorithm, which integrates cognitive testing scores from self-respondents and proxy assessments when direct testing is unavailable and classifies participants into normal cognition, cognitive impairment, or dementia based on established thresholds.^14^ Disability was defined as the onset of functional impairment in activities of daily living (ADLs) and was considered present when participants reported difficulty or dependence in one or more core ADLs, including bathing, dressing, eating, transferring, toileting, or walking across a room.^14^ Death was determined through linkage with the National Death Index and other HRS follow-up procedures used to ascertain vital status.^14^ For the composite outcome, time to event was defined as time from baseline to the earliest occurrence of incident dementia, incident disability, or death; participants who did not experience an event were censored at their last available follow-up.

### Statistical analysis

Baseline characteristics were summarized across polygenic profile categories using descriptive statistics. Time-to-event analyses evaluated associations of polygenic profile with the composite outcome and each component outcome. Kaplan-Meier curves were constructed to estimate cumulative incidence across polygenic profile groups, and differences between groups were assessed using log-rank tests. To visualize how 31-year cumulative incidence varied across iPRS-DEM percentiles, cumulative incidence was estimated separately at each percentile and the percentile-specific estimates were smoothed using locally estimated scatterplot smoothing (LOESS). This analysis was repeated according to APOE ε4 carrier status.

Associations between polygenic profile and outcomes were examined using Cox proportional hazards regression models, with time since baseline as the time scale. Hazard ratios (HRs) and 95% confidence intervals (CIs) were estimated for intermediate and poor polygenic profiles using the favorable profile as the reference category. Model 1 adjusted for age, sex, genetic ancestry, and the first 10 genetic principal components. Model 2 additionally adjusted for hypertension, diabetes mellitus, and smoking history. Model 3 further adjusted for years of education and baseline frequent vigorous physical activity. The proportional hazards assumption was assessed using scaled Schoenfeld residual plots and was considered adequately satisfied. In a time-stratified sensitivity analysis, associations remained directionally consistent across follow-up periods.

In secondary analyses, models were repeated separately among participants of European and African ancestry. For these analyses, polygenic profile categories were defined using percentile cut points calculated separately within each ancestry group. Analyses were also stratified by APOE ε4 carrier status. Interaction between continuous standardized iPRS-DEM and APOE ε4 status was formally assessed using linear regression models for 31-year outcome risk, with the continuous score used to preserve information and maximize statistical power.

Polygenic scores were calculated in HRS genetic data using PLINK 2.0, based on previously published variant weights for iPRS-DEM.^25^ All statistical analyses were performed using R version 4.5.1 (R Foundation for Statistical Computing, Vienna, Austria).^26^

## RESULTS

### Analytic Sample

Of 45,234 participants enrolled in the Health and Retirement Study, 15,620 provided DNA samples, of whom 15,565 had genotype data after quality control and imputation. After excluding individuals with prevalent dementia or disability at baseline, 14,333 participants were included in the primary analysis. Participants were categorized into three polygenic profile groups: favorable (n=2,867), intermediate (n=8,600), and poor (n=2,866). The median age at baseline was 55 years, and 58% of participants were female. Baseline characteristics of the analytic sample are shown in Table 1. Cardiovascular risk factors, including hypertension and diabetes, were more prevalent among participants with a poor polygenic profile.

**Table 1.** Baseline characteristics of participants by polygenic profile category.

| *Cohort / characteristic* | *Favorable* | *Intermediate* | *Poor* |
| --- | --- | --- | --- |
| *All participants* | N = 2,867 | N = 8,600 | N = 2,866 |
| *Age at baseline, median (IQR), years* | 55 (52, 62) | 55 (52, 61) | 54 (51, 58) |
| *Female sex, n (%)* | 1,603 (56%) | 4,937 (57%) | 1,783 (62%) |
| *Hypertension, n (%)* | 859 (30%) | 2,435 (28%) | 1,275 (44%) |
| *Diabetes mellitus, n (%)* | 160 (5.6%) | 628 (7.3%) | 403 (14%) |
| *Smoking history, n (%)* | 1,455 (51%) | 4,884 (57%) | 1,676 (58%) |
| *Heart disease, n (%)* | 223 (7.8%) | 768 (8.9%) | 252 (8.8%) |
| *Stroke, n (%)* | 46 (1.6%) | 123 (1.4%) | 85 (3.0%) |
| *Education, median (IQR), years* | 13 (12, 16) | 12 (12, 15) | 12 (11, 14) |
| *Frequent vigorous physical activity at baseline, n (%)* | 869 (34%) | 2,511 (32%) | 750 (28%) |
| *European ancestry* | N = 2,079 | N = 6,234 | N = 2,079 |
| *Age at baseline, median (IQR), years* | 55 (52, 64) | 55 (52, 61) | 55 (52, 61) |
| *Female sex, n (%)* | 1,139 (55%) | 3,581 (57%) | 1,220 (59%) |
| *Hypertension, n (%)* | 631 (30%) | 1,783 (29%) | 588 (28%) |
| *Diabetes mellitus, n (%)* | 101 (4.9%) | 338 (5.4%) | 196 (9.4%) |
| *Smoking history, n (%)* | 1,074 (52%) | 3,510 (56%) | 1,290 (62%) |
| *Heart disease, n (%)* | 176 (8.5%) | 567 (9.1%) | 222 (11%) |
| *Stroke, n (%)* | 37 (1.8%) | 97 (1.6%) | 37 (1.8%) |
| *African ancestry* | N = 427 | N = 1,278 | N = 427 |
| *Age at baseline, median (IQR), years* | 55 (52, 59) | 54 (52, 58) | 54 (52, 58) |
| *Female sex, n (%)* | 245 (57%) | 812 (64%) | 265 (62%) |
| *Hypertension, n (%)* | 201 (47%) | 632 (49%) | 208 (49%) |
| *Diabetes mellitus, n (%)* | 46 (11%) | 166 (13%) | 79 (19%) |
| *Smoking history, n (%)* | 248 (58%) | 747 (58%) | 246 (58%) |
| *Heart disease, n (%)* | 32 (7.5%) | 113 (8.8%) | 33 (7.7%) |
| *Stroke, n (%)* | 10 (2.3%) | 39 (3.1%) | 14 (3.3%) |

*Overall polygenic profile categories were defined using cohort-wide 20th and 80th percentile cut points, whereas ancestry-specific categories were recalculated using within-ancestry percentile cut points. Therefore, ancestry-specific favorable, intermediate, and poor groups are not subsets of the corresponding overall groups, and category-specific counts are not expected to sum to the overall category counts. Values are median (IQR), n (%), or n as indicated.*

Over a median follow-up of 19 years (IQR 13-25), contributing 276,135 person-years, 7,023 participants experienced the composite outcome; the first event was dementia in 1,469, disability in 1,989, and death in 3,565 participants. Crude outcome frequencies by polygenic profile are shown in Table 2. In ancestry-stratified analyses, 10,392 participants were of European ancestry and 2,132 were of African ancestry. Baseline demographic characteristics were broadly similar across polygenic profile categories within each ancestry group, although cardiovascular risk factors such as hypertension and diabetes were more prevalent among participants of African ancestry.

**Table 2.** Composite and individual outcomes by polygenic profile category.

| *Cohort / outcome* | *Favorable* | *Intermediate* | *Poor* |
| --- | --- | --- | --- |
| *All participants* | N = 2,867 | N = 8,600 | N = 2,866 |
| *Composite outcome* | 1,327 (46.3%) | 4,196 (48.8%) | 1,500 (52.3%) |
| *Dementia* | 252 (8.8%) | 879 (10.2%) | 634 (22.1%) |
| *Disability* | 373 (13.0%) | 1,336 (15.5%) | 525 (18.3%) |
| *Death* | 1,163 (40.6%) | 3,544 (41.2%) | 1,079 (37.6%) |
| *European ancestry* | N = 2,079 | N = 6,234 | N = 2,079 |
| *Composite outcome* | 996 (47.9%) | 3,038 (48.7%) | 1,079 (51.9%) |
| *Dementia* | 175 (8.4%) | 564 (9.0%) | 200 (9.6%) |
| *Disability* | 267 (12.8%) | 923 (14.8%) | 349 (16.8%) |
| *Death* | 891 (42.9%) | 2,657 (42.6%) | 918 (44.2%) |
| *African ancestry* | N = 427 | N = 1,278 | N = 427 |
| *Composite outcome* | 236 (55.3%) | 636 (49.8%) | 252 (59.0%) |
| *Dementia* | 104 (24.4%) | 312 (24.4%) | 106 (24.8%) |
| *Disability* | 85 (19.9%) | 217 (17.0%) | 85 (19.9%) |
| *Death* | 172 (40.3%) | 442 (34.6%) | 182 (42.6%) |

*Overall polygenic profile categories were defined using cohort-wide 20th and 80th percentile cut points, whereas ancestry-specific categories were recalculated using within-ancestry percentile cut points.*
Therefore, ancestry-specific favorable, intermediate, and poor groups are not subsets of the corresponding overall groups, and are not expected to sum to the overall.

### Polygenic profile and survival free of dementia and disability

Polygenic profile was associated with an increased risk of the composite outcome of dementia, disability, or death. Cumulative incidence curves demonstrated progressive separation across polygenic profile categories, with the highest cumulative incidence observed among participants with a poor polygenic profile (log-rank P<0.001; Figure 1A). When iPRS-DEM was examined across its full percentile distribution, 31-year cumulative incidence increased from 52% at the 1st percentile to 79% at the 100th percentile (Figure 2A).

**Figure 1.**
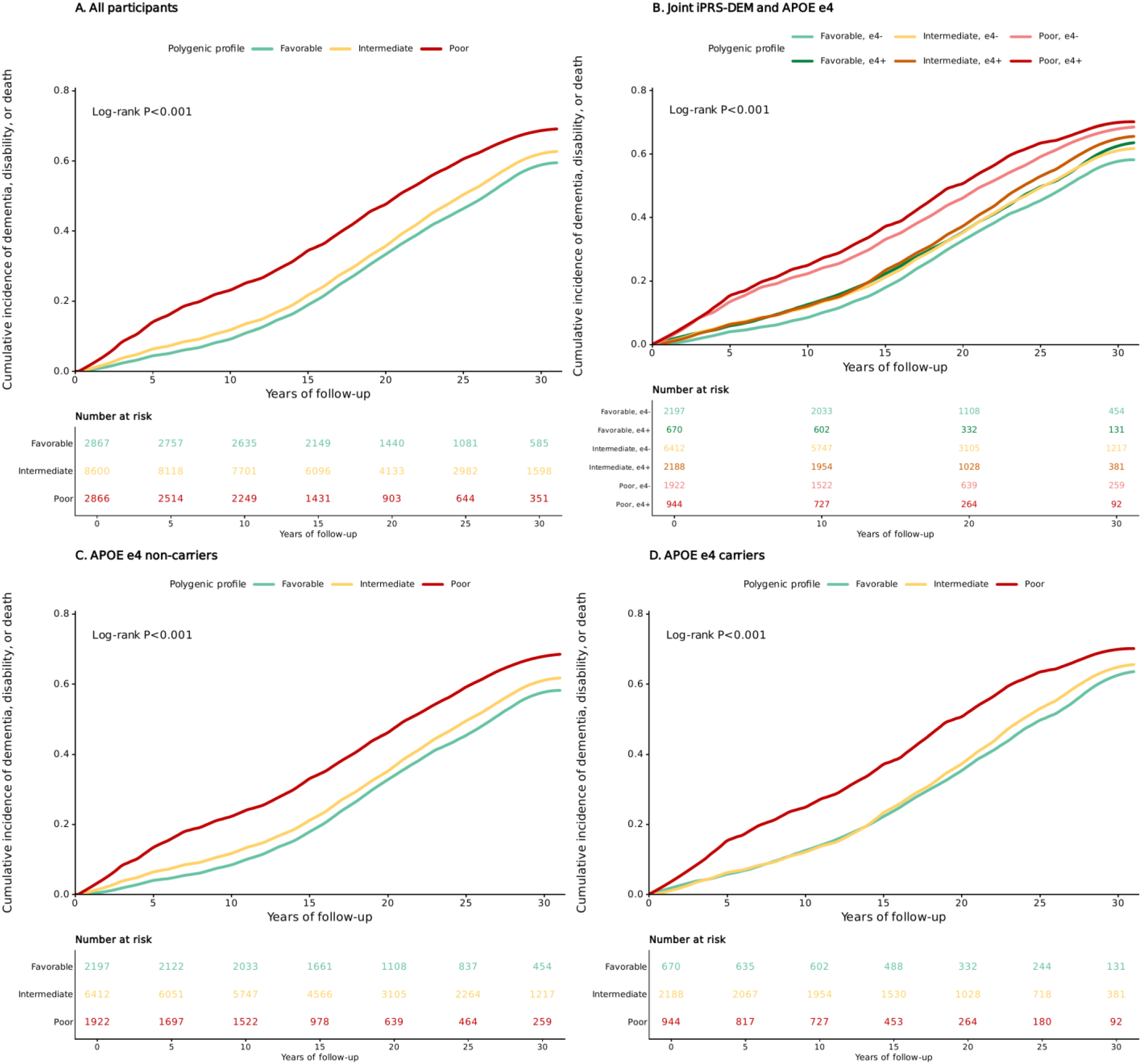
Cumulative incidence of dementia, disability, or death according to iPRS-DEM polygenic profile and APOE ε4 carrier status. (A) Cumulative incidence according to favorable, intermediate, and poor polygenic profiles in the overall analytic sample. (B) Cumulative incidence across six joint iPRS-DEM/APOE ε4 groups. (C) Cumulative incidence according to polygenic profile among APOE ε4 non-carriers. (D) Cumulative incidence according to polygenic profile among APOE ε4 carriers. Numbers at risk are shown below each panel; P values are from log-rank tests. APOE, apolipoprotein E; iPRS-DEM, integrated polygenic risk score for dementia; e4-, APOE ε4 non-carrier; e4+, APOE ε4 carrier.

**Figure 2.**
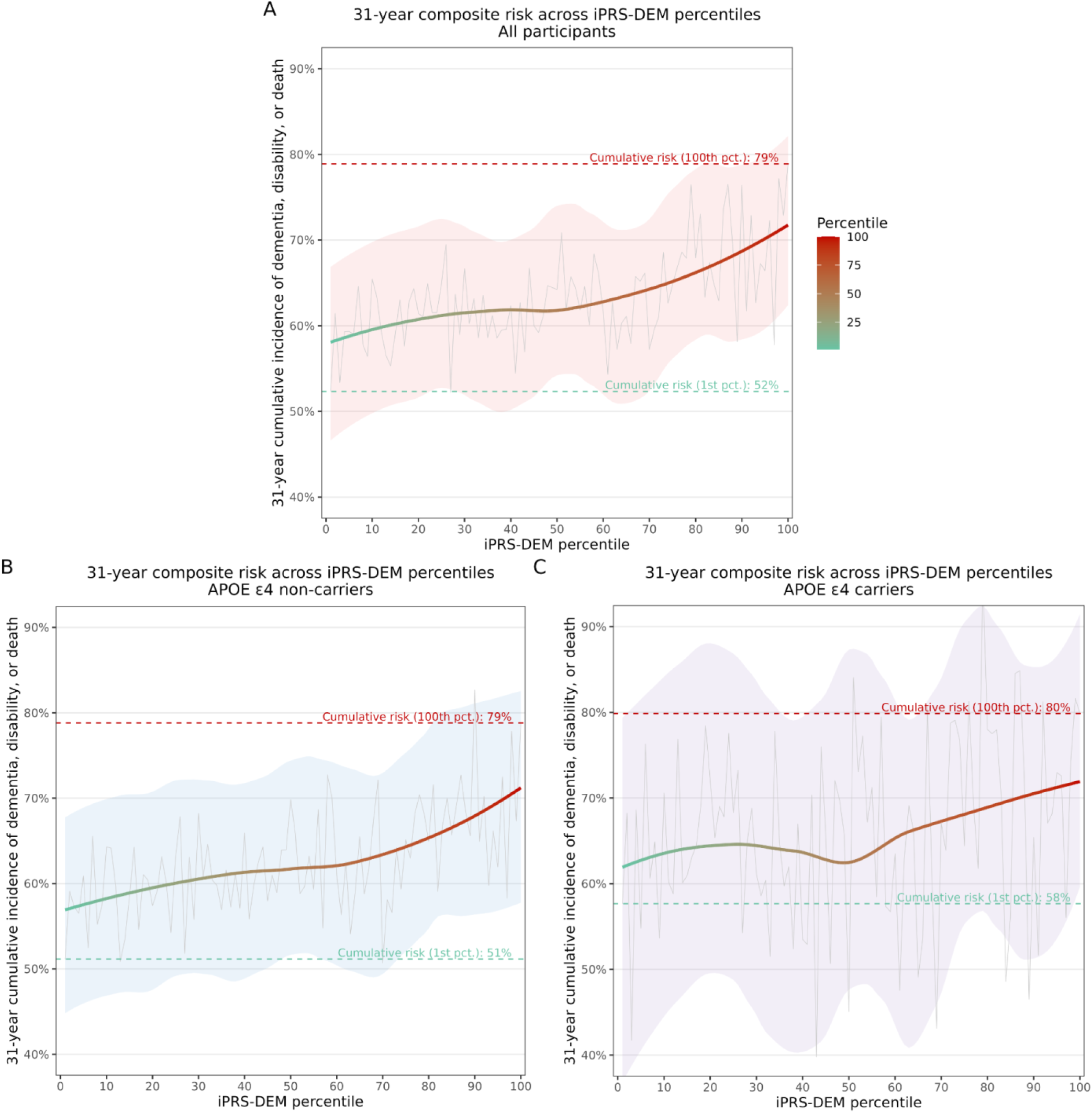
Thirty-one-year cumulative incidence of dementia, disability, or death across iPRS-DEM percentiles. (A) Overall analytic sample. (B) APOE ε4 non-carriers. (C) APOE ε4 carriers. Cumulative incidence was estimated separately at each iPRS-DEM percentile, and percentile-specific estimates and confidence intervals were smoothed using locally estimated scatterplot smoothing (LOESS) for visualization. APOE, apolipoprotein E; iPRS-DEM, integrated polygenic risk score for dementia; LOESS, locally estimated scatterplot smoothing.

In Cox proportional hazards models adjusted for age, sex, genetic ancestry, and genetic principal components, participants in the intermediate polygenic profile group had a higher risk of the composite outcome compared with those in the favorable profile group (HR 1.14, 95% CI 1.07-1.21), while those in the poor polygenic profile group had a higher risk (HR 1.43, 95% CI 1.26-1.63; Table 3A). When evaluating the individual components of the composite outcome, the relative association with polygenic profile was strongest for dementia. For poor versus favorable polygenic profiles, the HR was 1.58 (95% CI 1.21-2.05) for dementia, 1.43 (95% CI 1.14-1.79) for disability, and 1.33 (95% CI 1.15-1.54) for death (Table 3A). Associations were similar after additional adjustment for cardiovascular risk factors and after further adjustment for education and physical activity (Table 3A).

**Table 3.**
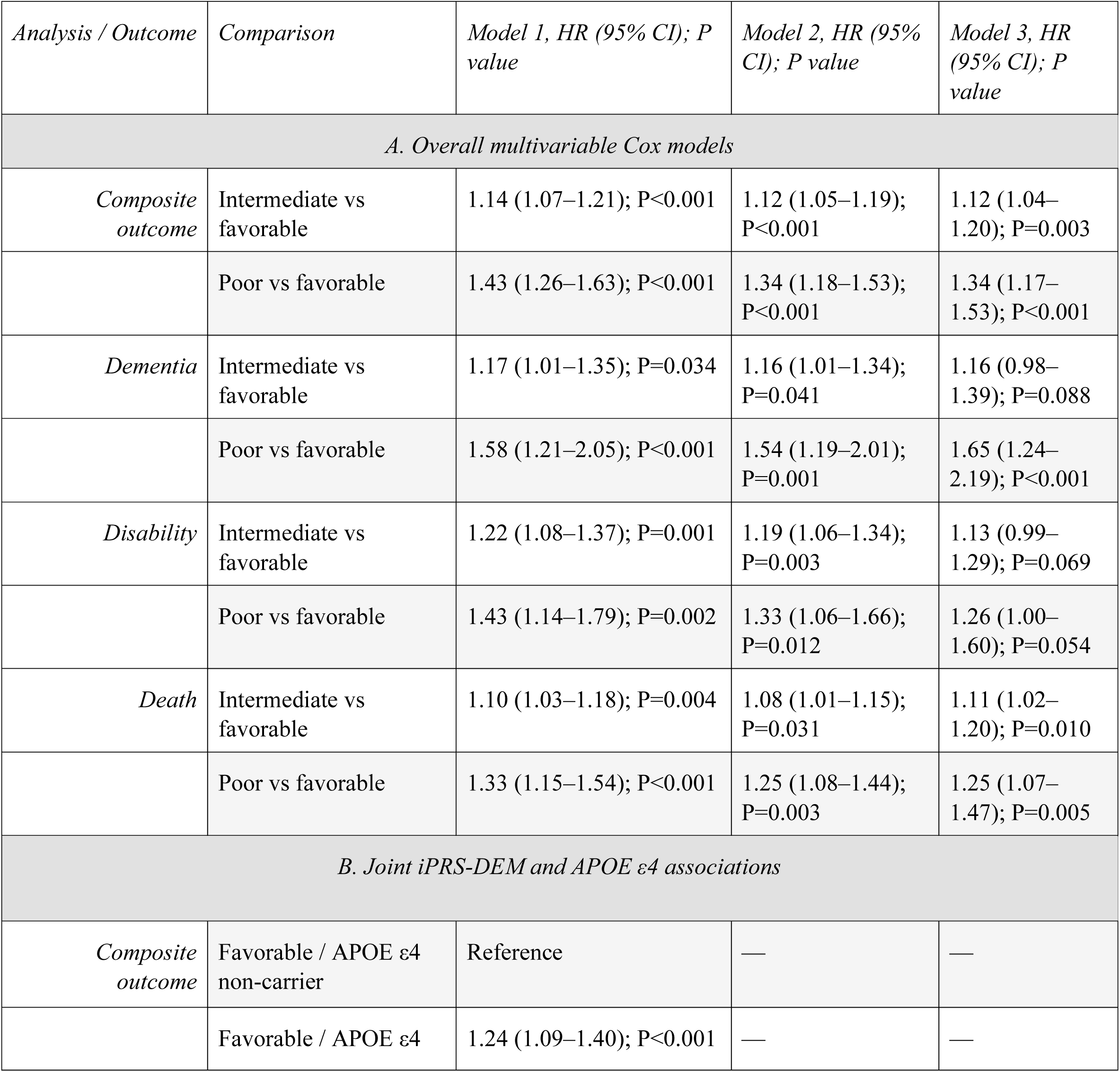
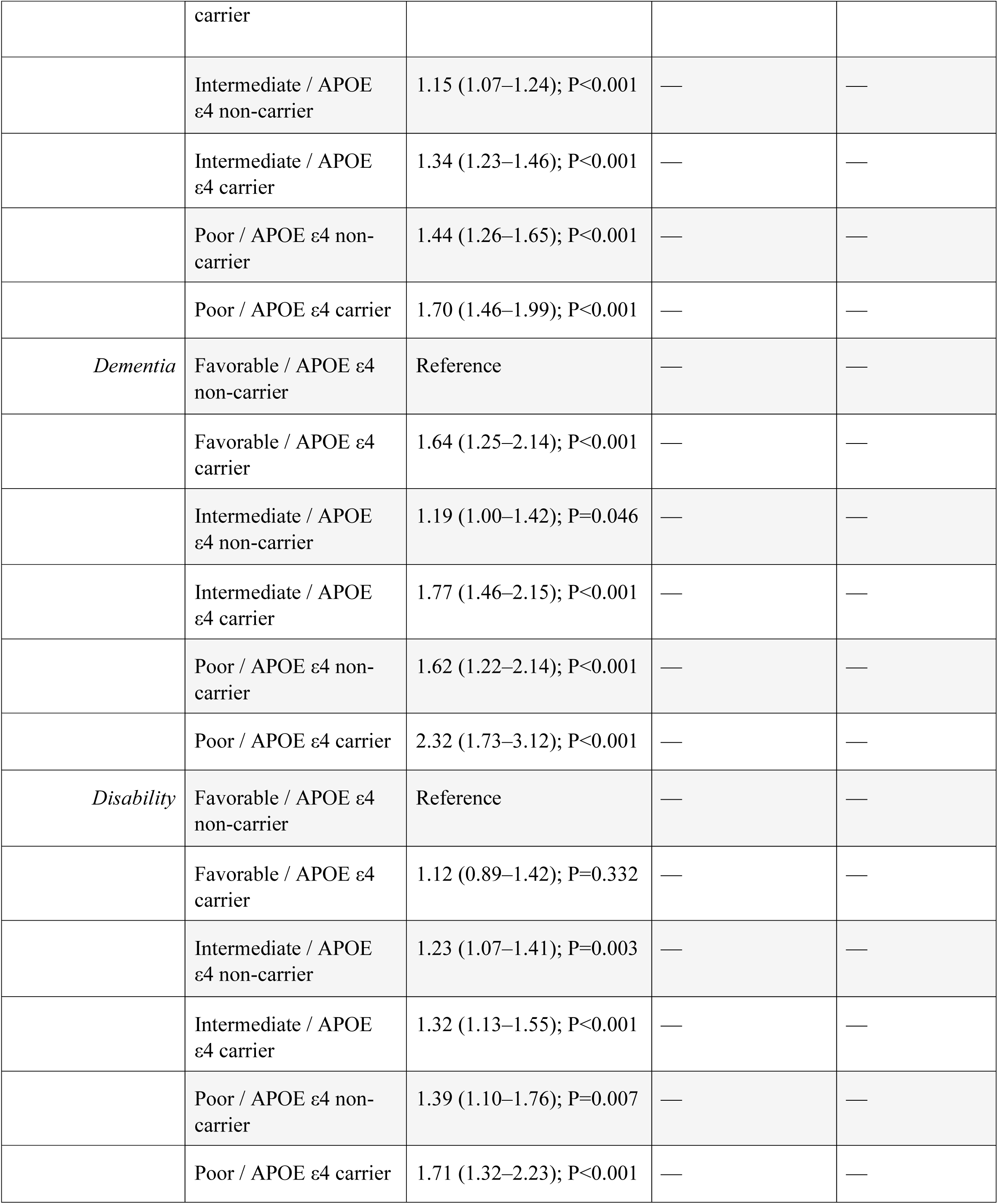
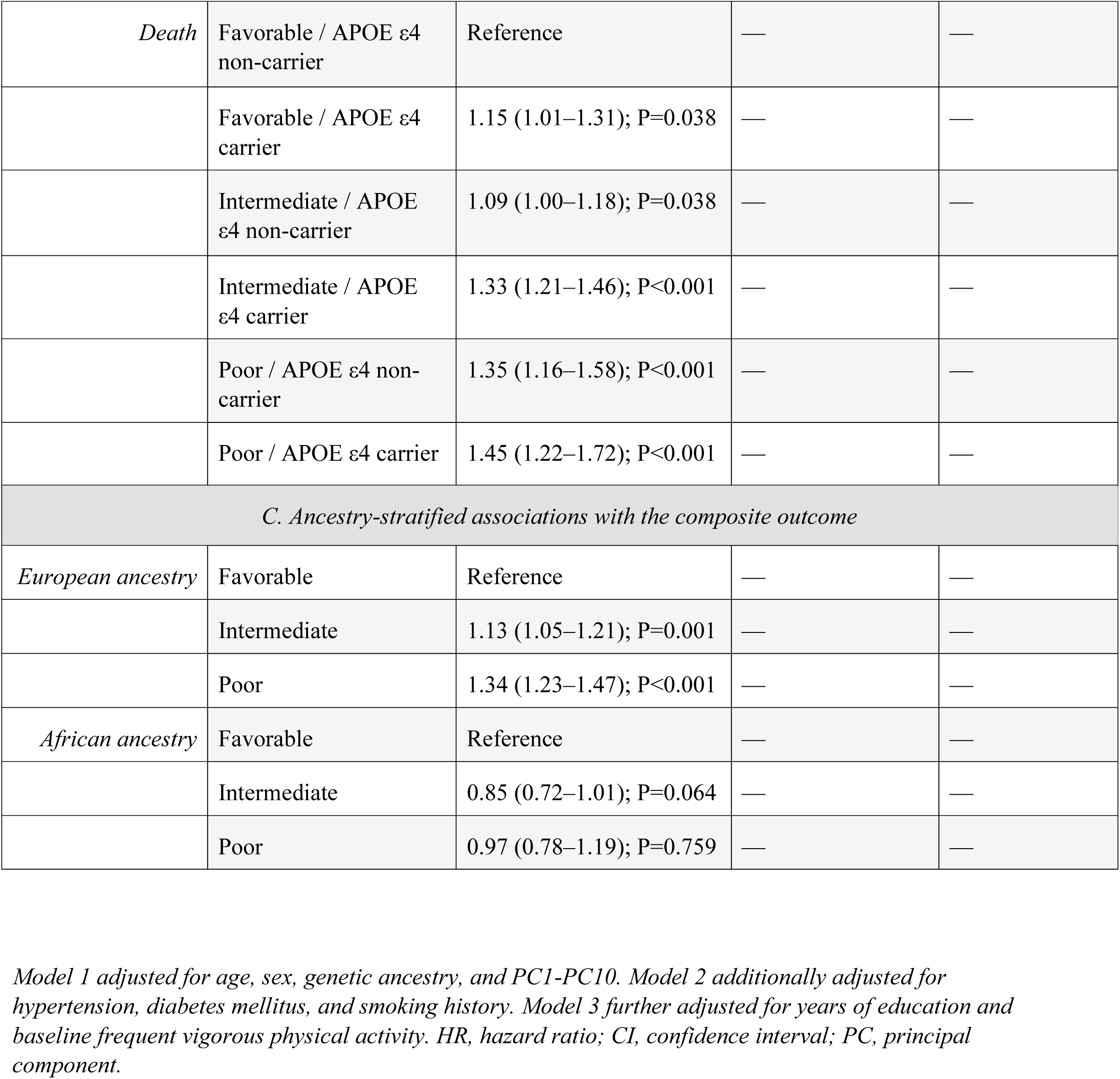
Multivariable associations of polygenic profile and APOE ε4 status with dementia, disability, or death.

### Stratified analyses and interaction with APOE ε4

We next evaluated whether the association between polygenic profile and outcomes differed according to APOE ε4 carrier status. Cumulative incidence of the composite outcome increased across joint iPRS-DEM/APOE ε4 groups, with the highest cumulative incidence observed among participants with both a poor polygenic profile and APOE ε4 carrier status (log-rank P<0.001; Figure 1B). Progressive separation across polygenic profile categories was also observed separately among APOE ε4 non-carriers and carriers (Figure 1C-D). The graded pattern in 31-year composite cumulative incidence was observed within both APOE ε4 strata, increasing from 51% to 79% across the 1st to 100th iPRS-DEM percentiles among non-carriers and from 58% to 80% among carriers (Figure 2B-C).

In adjusted joint Cox models, compared with participants with a favorable polygenic profile who were APOE ε4 non-carriers, the hazard of the composite outcome was higher among those with a poor profile who were APOE ε4 non-carriers (HR 1.44, 95% CI 1.26-1.65) and was highest among those with a poor profile who were APOE ε4 carriers (HR 1.70, 95% CI 1.46-1.99; Table 3B).

An interaction between continuous iPRS-DEM and APOE ε4 status was observed for 31-year dementia risk (P=0.016), whereas no evidence of interaction was observed for the composite outcome, disability, or death (Supplementary Table 1). For dementia, relative to participants with a favorable polygenic profile who were APOE ε4 non-carriers, the HR was 1.62 (95% CI 1.22-2.14) among those with a poor profile who were APOE ε4 non-carriers and 2.32 (95% CI 1.73-3.12) among those with a poor profile who were APOE ε4 carriers (Table 3B).

### Ancestry-stratified analyses

Among participants of European ancestry, cumulative incidence of the composite outcome increased across polygenic profile categories, with the highest cumulative incidence observed among participants with a poor polygenic profile (log-rank P<0.001; Supplementary Figure 4). Results were consistent with those in the overall cohort: compared with participants with a favorable polygenic profile, the adjusted HR for the composite outcome was 1.13 (95% CI 1.05-1.21) for the intermediate profile and 1.34 (95% CI 1.23-1.47) for the poor profile (Table 3C).

Among participants of African ancestry, cumulative incidence differed across polygenic profile categories in unadjusted analyses (log-rank P=0.002; Supplementary Figure 5); however, adjusted associations with the composite outcome were not observed. Compared with the favorable polygenic profile, the HR was 0.85 (95% CI 0.72-1.01) for the intermediate profile and 0.97 (95% CI 0.78-1.19) for the poor profile (Table 3C).

## DISCUSSION

In this study, we evaluated the association between a polygenic profile for dementia and survival free of dementia and disability in a large, nationally representative cohort of middle-aged and older adults from the Health and Retirement Study. We found that individuals with intermediate and poor polygenic profiles had progressively higher risk of the composite outcome compared with those with a favorable profile. The relative association was strongest for dementia, followed by disability and death. The highest dementia risk was observed among participants with both a poor polygenic profile and APOE ε4 carrier status, and an interaction between continuous iPRS-DEM and APOE ε4 status was observed for 31-year dementia risk. Results were consistent among participants of European ancestry, whereas adjusted associations were not observed among participants of African ancestry.

Prior studies have consistently shown that polygenic risk scores are associated with increased risk of Alzheimer disease and dementia.^18–20^ For example, Desikan et al.^18^ demonstrated that a polygenic hazard score substantially modified age at onset of Alzheimer disease, even among individuals without APOE ε4, highlighting the contribution of polygenic risk beyond single-gene effects. Similarly, in a prospective community-based cohort, Stocker et al.^21^ found that an Alzheimer disease polygenic risk score was associated with higher risk of incident dementia and improved risk prediction beyond traditional demographic factors. In addition, large population-based analyses such as UK Biobank have shown that higher polygenic risk is associated with significantly increased incidence of dementia in older adults.^20,22^ However, these studies have primarily focused on dementia as a single outcome and have not assessed associations with composite aging outcomes. Importantly, they have not incorporated disability, a critical outcome in older adults with major implications for independence, quality of life, and healthcare utilization. As a result, the broader impact of polygenic susceptibility on aging-related outcomes remains incompletely characterized.

To address this gap, we used the HRS to test whether dementia-related polygenic burden is associated with maintenance of functionally meaningful health in later life. Rather than evaluating dementia alone, we focused on a composite outcome of dementia, disability, or death, reflecting outcomes that are highly relevant to aging populations. We found a graded association between polygenic profile and this composite outcome, with progressively higher risk among participants in the intermediate and poor polygenic profile groups. The relative association was strongest for dementia, but significant associations were also observed for disability and death. These findings suggest that polygenic profiling for dementia may capture risk relevant to broader aging-related health trajectories rather than cognitive outcomes alone.

One important implication of these findings is that polygenic susceptibility measured from midlife remains associated with health trajectories extending into later life. Participants entered the study at a median age of 55 years and were followed for a median of 19 years, allowing genetic associations to be evaluated across a substantial period of aging. Our findings suggest that polygenic susceptibility remains relevant to clinically meaningful aging-related outcomes over long-term follow-up. This is consistent with broader evidence that genetic factors contribute to healthy aging and longevity, even though aging trajectories are also strongly shaped by lifestyle, social, and environmental exposures.^23,24^

Another important implication of these findings is that polygenic profiling may help identify subgroups at higher risk for adverse aging-related outcomes. While polygenic scores have previously been shown to improve dementia risk stratification, our findings extend this work by demonstrating their relevance to composite outcomes that incorporate functional decline and survival. In this context, polygenic profile further stratified risk in conjunction with APOE ε4 status, with the highest dementia risk observed among individuals with both a poor polygenic profile and APOE ε4 carrier status. These results suggest that polygenic profiling may help identify individuals at particularly high risk who could be prioritized for preventive strategies or considered for enrichment in clinical trials targeting dementia and functional decline. However, clinical application requires caution, particularly across ancestries. Associations observed in the overall and European-ancestry analyses were not reproduced among participants of African ancestry, consistent with recognized limitations in the cross-ancestry transferability of current polygenic scores.^10–12^ More diverse genomic discovery studies and ancestry-informed approaches will be needed before such tools can be broadly applied.

This study has several limitations. First, dementia was ascertained using a validated HRS cognitive classification algorithm, whereas disability was based on self- or proxy-reported difficulty with activities of daily living; both measures remain subject to potential outcome misclassification. Second, iPRS-DEM was developed primarily for dementia prediction and was not specifically optimized to predict disability or mortality; associations with these outcomes should therefore be interpreted cautiously. Third, the smaller African-ancestry sample and the known ancestry dependence of polygenic-score performance limit conclusions regarding generalizability across populations. Finally, as an observational genetic association study, these findings do not establish causality and should not be interpreted as evidence that polygenic profiling alone is sufficient to guide clinical decision-making.

In conclusion, polygenic profiling for dementia was associated with the composite risk of dementia, disability, or death, with the strongest association observed for dementia. Joint consideration of iPRS-DEM and APOE ε4 further identified individuals at elevated dementia risk, including evidence of interaction for 31-year dementia risk. These findings support the potential value of polygenic profiling for risk stratification while highlighting important limitations related to ancestry, generalizability, and clinical translation.

## Acknowledgments

The authors thank the participants and staff of the Health and Retirement Study for their longstanding contributions to aging research. The Health and Retirement Study is sponsored by the National Institute on Aging (U01AG009740) and conducted by the University of Michigan.

## Data Availability

The HRS (Health and Retirement Study) is sponsored by the National Institute on Aging (NIA U01AG009740 and NIA R01AG073289) and is conducted by the University of Michigan. Data used in this study are available through the HRS. The iPRS-DEM score weights are publicly available through the Polygenic Score Catalog (PGS005170).

## REFERENCES

1. McNeil JJ, Woods RL, Nelson MR, et al. Effect of Aspirin on Disability-free Survival in the Healthy Elderly. N Engl J Med. 2018;379(16):1499–1508. doi:10.1056/NEJMoa1800722

2. Zoungas S, Curtis A, Spark S, et al. Statins for extension of disability-free survival and primary prevention of cardiovascular events among older people: protocol for a randomised controlled trial in primary care (STAREE trial). BMJ Open. 2023;13(4):e069915. doi:10.1136/bmjopen-2022-069915

3. Duke University. PRagmatic EValuation of evENTs And Benefits of Lipid-Lowering in oldEr Adults (PREVENTABLE). ClinicalTrials.gov; 2026. Accessed July 16, 2026. https://clinicaltrials.gov/study/NCT04262206

4. Corder EH, Saunders AM, Risch NJ, et al. Protective effect of apolipoprotein E type 2 allele for late onset Alzheimer disease. Nat Genet. 1994;7(2):180–184. doi:10.1038/ng0694-180

5. Corder EH, Saunders AM, Strittmatter WJ, et al. Gene Dose of Apolipoprotein E Type 4 Allele and the Risk of Alzheimer’s Disease in Late Onset Families. Science. 1993;261(5123):921–923. doi:10.1126/science.8346443

6. Bennet AM, Di Angelantonio E, Ye Z, et al. Association of Apolipoprotein E Genotypes With Lipid Levels and Coronary Risk. JAMA. 2007;298(11):1300–1311. doi:10.1001/jama.298.11.1300

7. Schächter F. Causes, Effects, and Constraints in the Genetics of Human Longevity. Am J Hum Genet. 1998;62(5):1008–1014. doi:10.1086/301849

8. Clocchiatti-Tuozzo S, Szejko N, Rivier CA, et al. APOE epsilon variants and composite risk of dementia, disability, and death in the Health and Retirement Study. doi:10.1111/jgs.19043

9. D’Aoust T, Clocchiatti-Tuozzo S, Rivier CA, et al. Polygenic score integrating neurodegenerative and vascular risk informs dementia risk stratification. Alzheimer’s & Dementia. 2025;21(3):e70014. doi:10.1002/alz.70014

10. Theoretical and empirical quantification of the accuracy of polygenic scores in ancestry divergent populations. Nat Commun. Accessed July 16, 2026. https://www.nature.com/articles/s41467-020-17719-y

11. Duncan L, Shen H, Gelaye B, et al. Analysis of polygenic risk score usage and performance in diverse human populations. Nat Commun. 2019;10(1):3328. doi:10.1038/s41467-019-11112-0

12. Clinical use of current polygenic risk scores may exacerbate health disparities. Nat Genet. Accessed July 16, 2026. https://www.nature.com/articles/s41588-019-0379-x

13. Sonnega A, Faul JD, Ofstedal MB, Langa KM, Phillips JW, Weir DR. Cohort Profile: the Health and Retirement Study (HRS). Int J Epidemiol. 2014;43(2):576–585. doi:10.1093/ije/dyu067

14. RAND HRS Longitudinal File 2022 (V1). Produced by the RAND Center for the Study of Aging, with funding from the National Institute on Aging and the Social Security Administration. Santa Monica, CA (May 2025).

15. Goldstein JI, Crenshaw A, Carey J, et al. zCall: a rare variant caller for array-based genotyping. Bioinformatics. 2012;28(19):2543–2545. doi:10.1093/bioinformatics/bts479

16. Delaneau O, Marchini J, Zagury JF. A linear complexity phasing method for thousands of genomes. Nat Methods. 2012;9(2):179–181. doi:10.1038/nmeth.1785

17. Howie BN, Donnelly P, Marchini J. A Flexible and Accurate Genotype Imputation Method for the Next Generation of Genome-Wide Association Studies. PLoS Genet. 2009;5(6):e1000529. doi:10.1371/journal.pgen.1000529

18. Desikan RS, Fan CC, Wang Y, et al. Genetic assessment of age-associated Alzheimer disease risk: Development and validation of a polygenic hazard score. PLoS Med. 2017;14(3):e1002258. doi:10.1371/journal.pmed.1002258

19. Escott-Price V, Sims R, Bannister C, et al. Common polygenic variation enhances risk prediction for Alzheimer’s disease. Brain. 2015;138(12):3673–3684. doi:10.1093/brain/awv268

20. Leonenko G, Baker E, Stevenson-Hoare J, et al. Identifying individuals with high risk of Alzheimer’s disease using polygenic risk scores. Nat Commun. 2021;12(1):4506. doi:10.1038/s41467-021-24082-z

21. Stocker H, Perna L, Weigl K, et al. Prediction of clinical diagnosis of Alzheimer’s disease, vascular, mixed, and all-cause dementia by a polygenic risk score and APOE status in a community-based cohort prospectively followed over 17 years. Mol Psychiatry. 2021;26(10):5812–5822. doi:10.1038/s41380-020-0764-y

22. Alzheimer’s disease polygenic risk’s association with all-cause dementia through the plasma metabolome in the UK Biobank study. GeroScience. Accessed July 16, 2026. https://link.springer.com/article/10.1007/s11357-025-01724-4

23. Deelen J, Evans DS, Arking DE, et al. A meta-analysis of genome-wide association studies identifies multiple longevity genes. Nat Commun. 2019;10(1):3669. doi:10.1038/s41467-019-11558-2

24. Timmers PR, Mounier N, Lall K, et al. Genomics of 1 million parent lifespans implicates novel pathways and common diseases and distinguishes survival chances. eLife. 2019;8:e39856. doi:10.7554/eLife.39856

25. Chang CC, Chow CC, Tellier LCAM, Vattikuti S, Purcell SM, Lee JJ. Second-generation PLINK: rising to the challenge of larger and richer datasets. GigaScience. 2015;4:7. doi:10.1186/s13742-015-0047-8

26. R Core Team. R: A Language and Environment for Statistical Computing. R Foundation for Statistical Computing; Vienna, Austria. https://www.R-project.org/

