## Supplementary Material for "Multi-trait Polygenic Profiling and Survival Free of Dementia and Disability: Results from the Health and Retirement Study"

**Supplementary Table 1. Interaction between continuous iPRS-DEM and APOE ε4 status by outcome**

| Outcome | Interaction β (95% CI) | P value |
|---|---|---|
| Composite | 0.005 (-0.017 to 0.026) | 0.674 |
| Dementia | 0.025 (0.005 to 0.045) | 0.016 |
| Disability | 0.014 (-0.005 to 0.032) | 0.142 |
| Death | 0.009 (-0.013 to 0.031) | 0.433 |

**Supplementary Table 2. Time-stratified sensitivity analysis for the composite outcome**

| Follow-up period | Contrast | HR (95% CI) | P value |
|---|---|---|---|
| 0-10 years | Intermediate vs Favorable | 1.25 (1.09-1.43) | 0.001 |
| 0-10 years | Poor vs Favorable | 1.67 (1.32-2.12) | <0.001 |
| >10-20 years | Intermediate vs Favorable | 1.07 (0.98-1.18) | 0.134 |
| >10-20 years | Poor vs Favorable | 1.29 (1.05-1.59) | 0.016 |
| >20 years | Intermediate vs Favorable | 1.15 (1.03-1.28) | 0.014 |
| >20 years | Poor vs Favorable | 1.43 (1.12-1.82) | 0.004 |

*Cox models adjusted for age, sex, genetic ancestry, and PC1-PC10. Favorable polygenic profile was the reference category. HR, hazard ratio; CI, confidence interval; PC, principal component.*

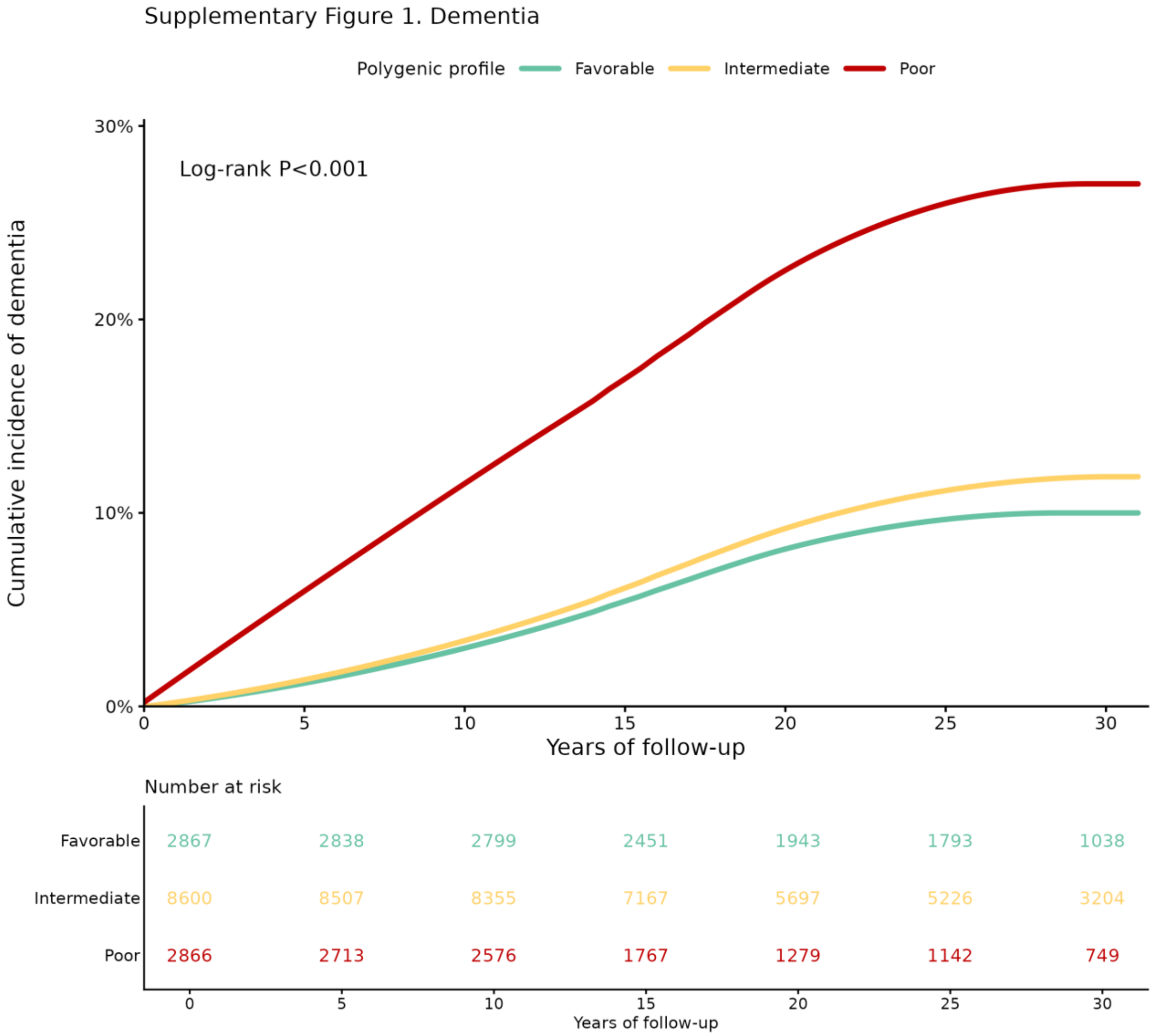


**Supplementary Figure 1.** Cumulative incidence of dementia according to iPRS-DEM polygenic profile. Numbers at risk are shown below the plot; P value is from the log-rank test.

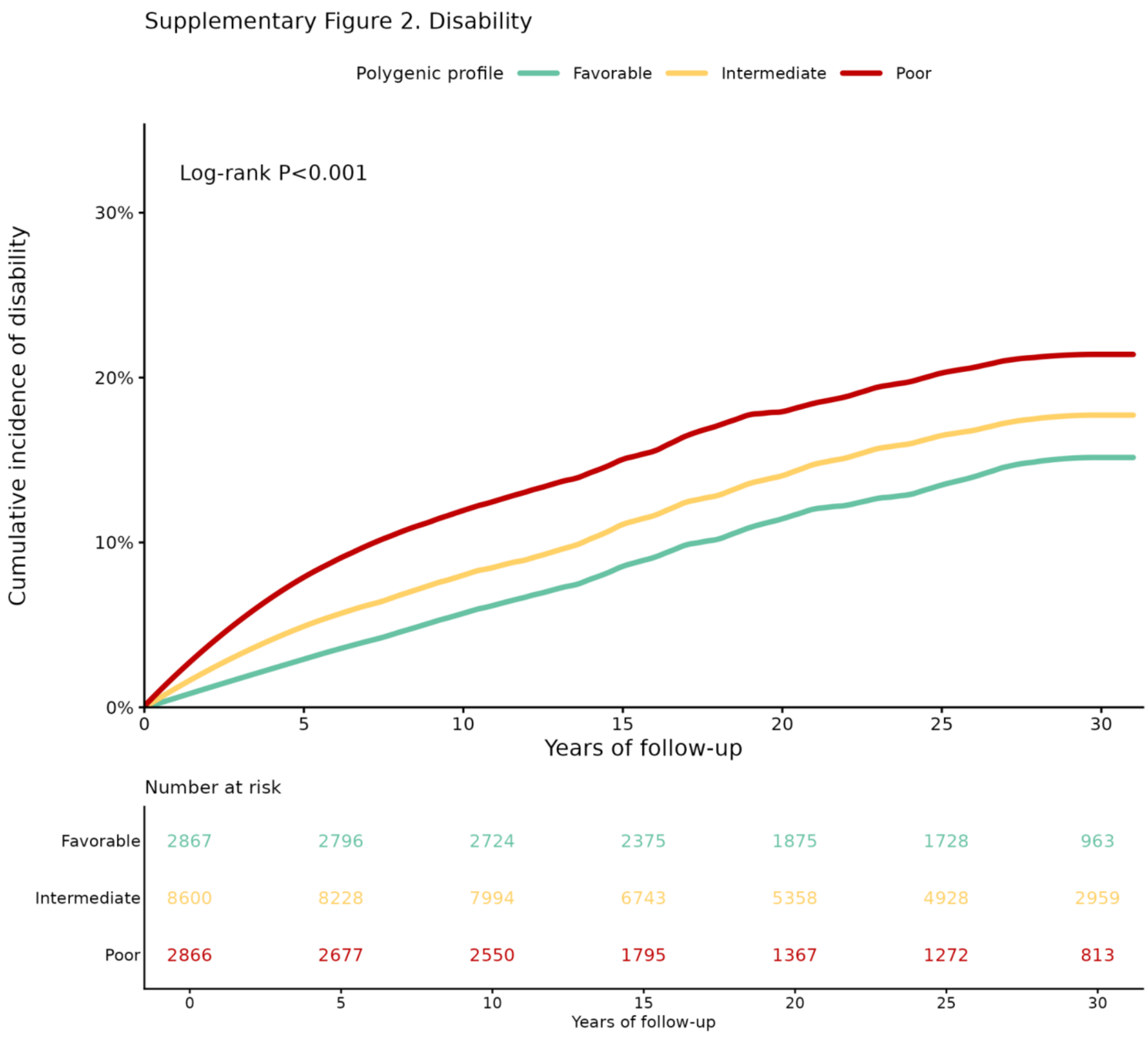


**Supplementary Figure 2.** Cumulative incidence of disability according to iPRS-DEM polygenic profile. Numbers at risk are shown below the plot; P value is from the log-rank test.

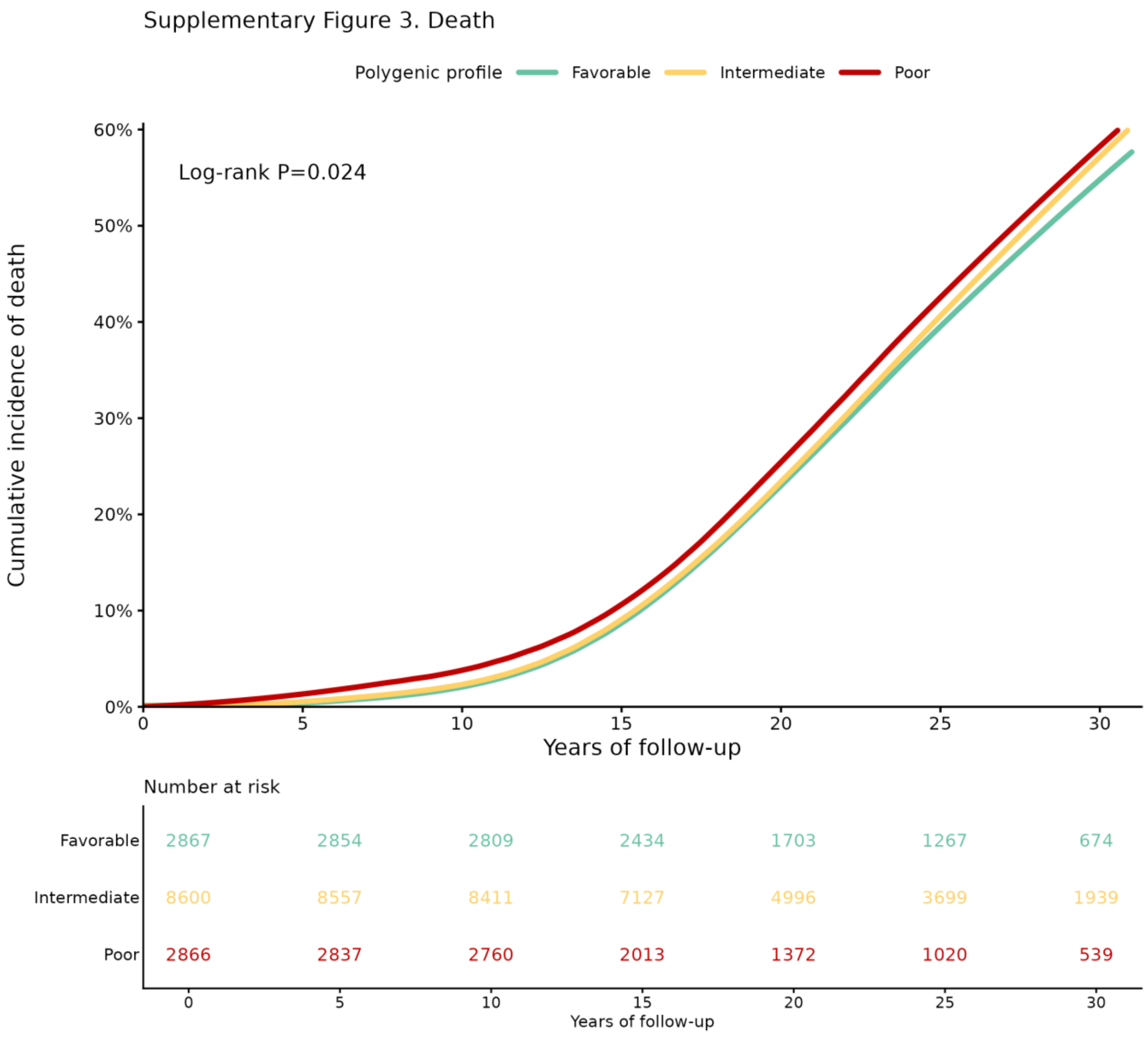


**Supplementary Figure 3.** Cumulative incidence of death according to iPRS-DEM polygenic profile. Numbers at risk are shown below the plot; P value is from the log-rank test.

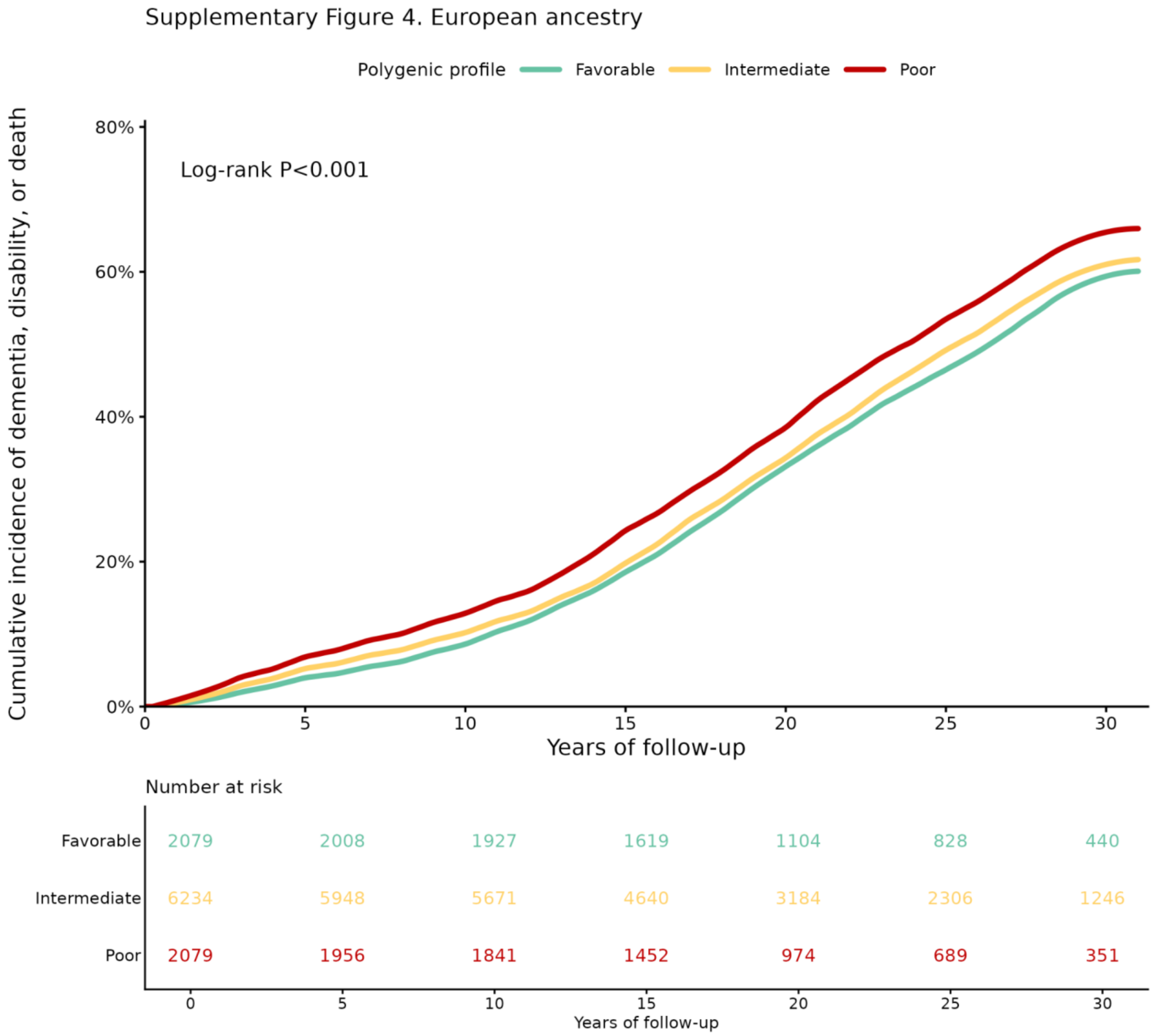


**Supplementary Figure 4.** Cumulative incidence of dementia, disability, or death according to iPRS-DEM polygenic profile among participants of European ancestry. Polygenic profile categories were defined using within-European-ancestry percentile cut points. Numbers at risk are shown below the plot; P value is from the log-rank test.

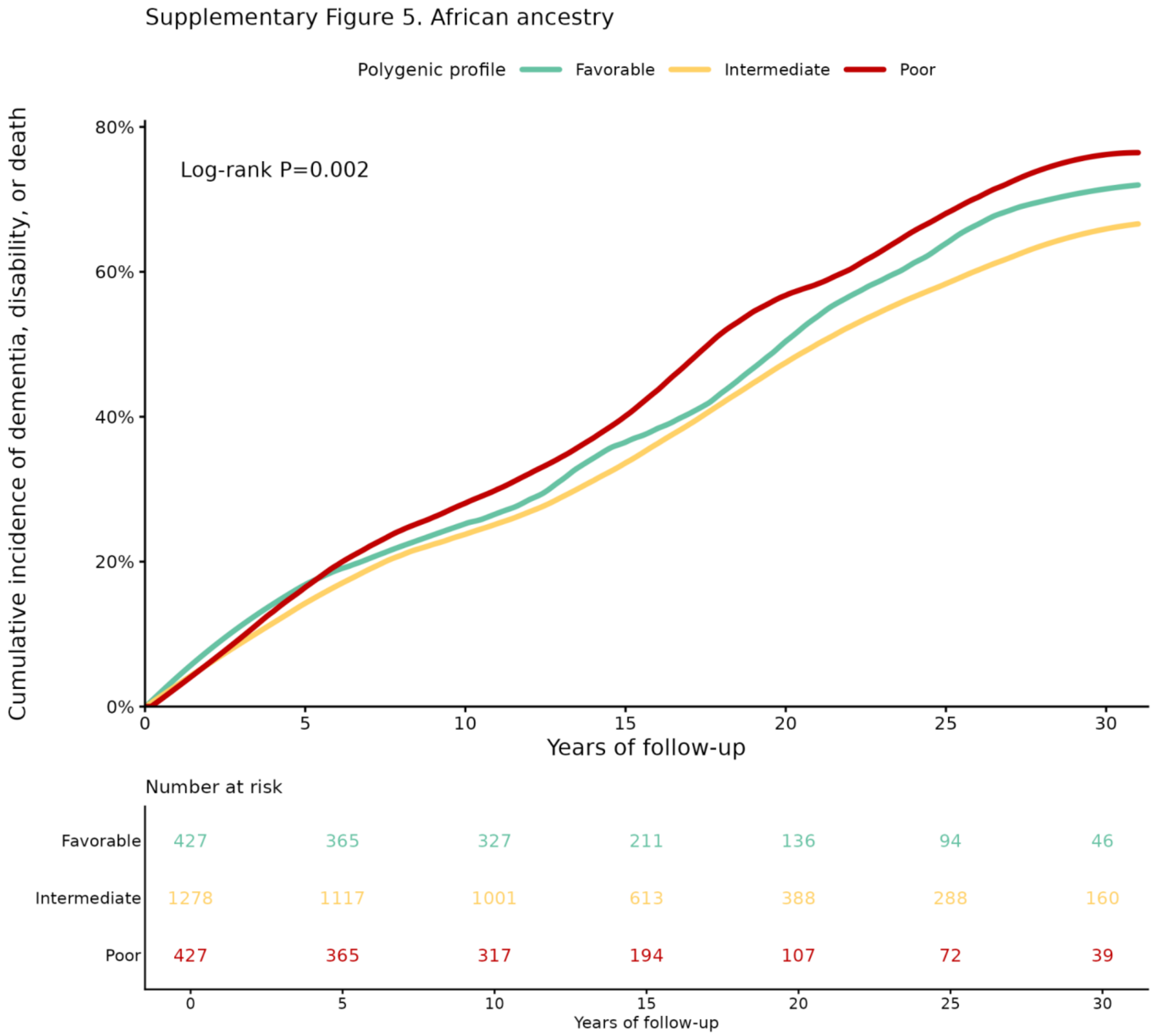


**Supplementary Figure 5.** Cumulative incidence of dementia, disability, or death according to iPRS-DEM polygenic profile among participants of African ancestry. Polygenic profile categories were defined using within-African-ancestry percentile cut points. Numbers at risk are shown below the plot; P value is from the log-rank test.

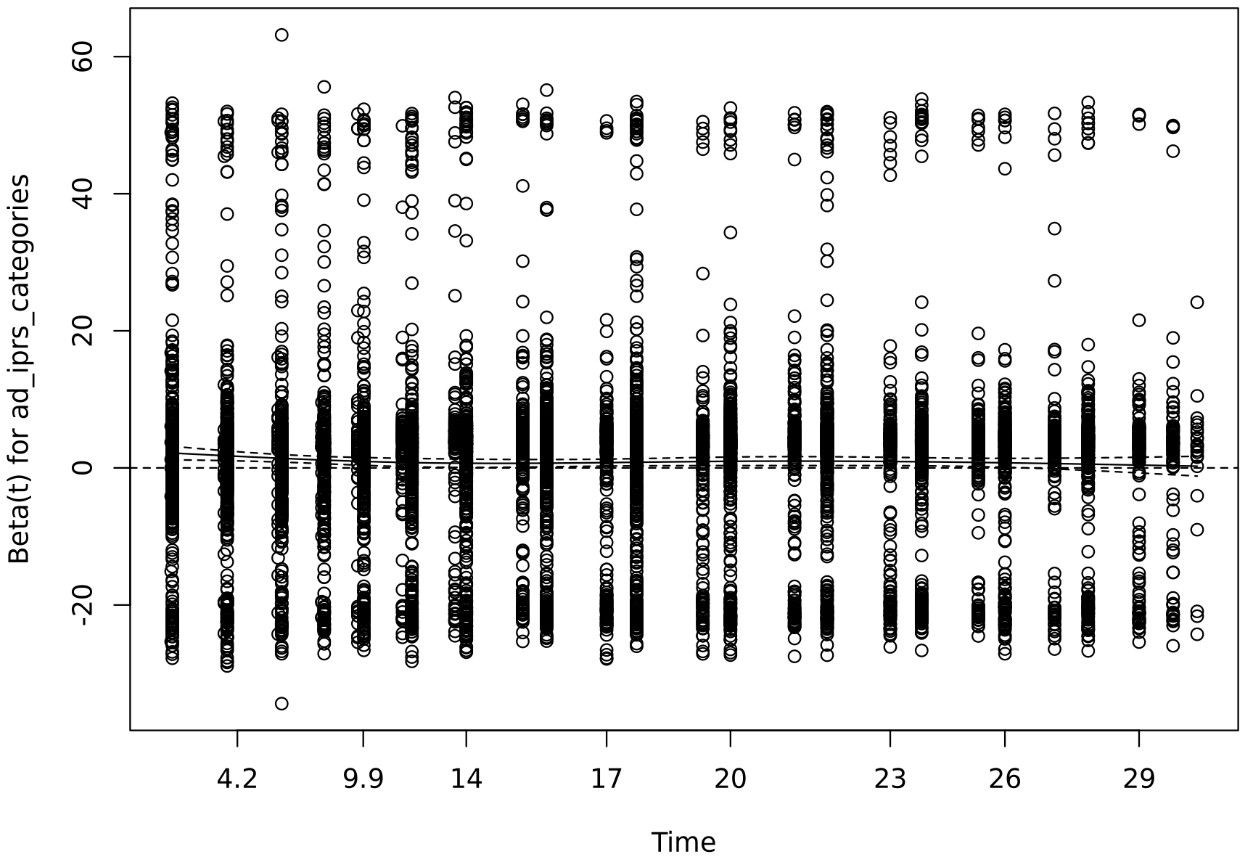


**Supplementary Figure 6.** Scaled Schoenfeld residual diagnostic for the primary polygenic-profile term in the composite-outcome Cox model. The horizontal reference line at zero and smoothed residual trend are shown to assess time-varying effects.

*Abbreviations: APOE, apolipoprotein E; iPRS-DEM, integrated polygenic risk score for dementia.*
